# Uncoded Clinical Features from Multilingual Electronic Health Records in Catalonia: Development and Validation Study

**DOI:** 10.64898/2026.09.16.26363030

**Authors:** Dan Ouchi, Silvia Riudavets, Alvaro Franquet, Silvia Fernández-García, Carl Llor, Ana Moragas-Moreno, Maria Giner-Soriano, Ferran Torres, Rosa Morros

## Abstract

**Background:** Unstructured free-text narratives in electronic health records (EHRs) contain critical clinical information that is not captured by structured standard codes. In Catalonia, primary care notes follow a semi-structured format known as MEAP (in Catalan). However, extracting structured variables using cloud-hosted commercial large language models (LLMs) raises substantial data privacy concerns and incurs high operational costs.

**Objective:** To develop and validate a privacy-preserving local hybrid pipeline combining a lightweight small language model (SLM) with deterministic regular expressions for extracting six uncoded urinary tract infection (UTI) clinical features (fever, nitrites, leukocytes, lumbar pain, abdominal pain, and haematuria) from primary care EHR narratives.

**Methods:** We deployed the open-weight model (Phi4-mini, 3.8B parameters) within the institutional firewall using Ollama. Prompts were iteratively refined in collaboration with clinicians, and regular expressions were applied post-inference. Validation was conducted across two independent arms: (1) a double-blind clinician gold standard review of 60 real patient records (consensus-resolved cases), and (2) an adversarial synthetic dataset of 720 notes enriched with linguistic noise, generated using GPT-4.1 and Grok-4.1. Point estimates and 95% confidence intervals (CIs) were computed for key performance metrics.

**Results:** The pipeline processed 15,498 MEAP narratives from a matched cohort of 2,962 primary care patients and identified 4,663 clinical feature occurrences. Patients progressing to acute pyelonephritis (cases) presented a higher features burden than non-progressing controls (71.3% vs. 55.8%; standardized mean difference [SMD] = 0.326), particularly for fever (33.0% vs. 9.0%; SMD = 0.616) and lumbar pain (29.0% vs. 9.8%; SMD = 0.502). In real-world clinician validation, overall performance yielded 93.8% accuracy (95% CI 90.6%-96.1%), 83.6% sensitivity (95% CI 73.0%-91.2%), 96.6% specificity (95% CI 93.6%-98.4%), 87.1% positive predictive value (PPV; 95% CI 77.0%-93.9%), and 95.5% negative predictive value (NPV; 95% CI 92.3%-97.7%). In synthetic stress-testing, the pipeline demonstrated 87.2% accuracy (95% CI 84.6%-89.6%), 73.9% sensitivity (95% CI 68.9%-78.5%), near-perfect specificity (99.5%, 95% CI 98.1%-99.9%), and 99.2% PPV (95% CI 97.2%-99.9%).

**Conclusions:** A privacy-preserving local hybrid framework combining a compact open-weight SLM with regular expression rules achieves high specificity and precision for extracting six uncoded clinical features from multilingual primary care narratives. Running entirely within institutional servers, this approach keeps patient data secure, meets privacy standards, and avoids Application Programming Interface (API) costs, making it a practical tool for EHR research, though with lower sensitivity for narratively complex descriptions.

## Introduction

Electronic Health Record (EHR) databases serve as indispensable resources for real-world epidemiological research and clinical predictive modelling [1], [2], [3], [4]. However, a substantial portion of critical clinical information remains locked in unstructured free-text clinical narratives rather than standardized diagnostic codes, such as International Classification of Diseases (ICD) or Anatomical Therapeutic Chemical [ATC] codes [5], [6]. This creates significant data opacity, as it omits granular clinical symptoms/signs, trajectories that are essential for assessing disease progression and treatment outcomes [7].

In primary care settings, clinical documentation often follows semi-structured narrative frameworks. Within the Information System for the Development of Research in Primary Care (SIDIAP) database in Catalonia, Spain, clinical notes are recorded using the MEAP format (*Motiu de consulta [reason for encounter], Exploració [physical examination findings], Avaluació [assessment], and Planificació [plan]*) [8], [9].

While MEAP records contain rich, real-time longitudinal data captured by primary care clinicians during patient encounters, extracting structured variables from these free-text narratives presents major computational and linguistic challenges [10], [11], [12]. Moreover, in regional healthcare systems like Catalonia, primary care narratives are multilingual (predominantly Catalan and Spanish), requiring extensive manual rule engineering [13].

The extraction of structured clinical variables from unstructured text has historically relied on rule-based Natural Language Processing (NLP) tools (e.g., cTAKES, MedSpacy) or fine-tuning BERT models [10], [14], [15], [16]. While highly specific, these methods suffer from poor adaptability to high linguistic variability, non-standard clinical abbreviations and unstructured narrative noise. Thus, in the recent years, large language models (LLMs) have been introduced as an effective alternative to automate the data extraction of important patient characteristics from electronic health records [17], [18].

Despite their efficacy, integrating commercial cloud-hosted LLM Application Programming Interface (APIs) (e.g., GPT-4) into clinical research introduces severe regulatory and ethical challenges. Sharing patient health information (PHI) with external cloud servers directly violates privacy frameworks and local health data governance protocols [19], [20]. In addition, high recurring API costs present scalability bottlenecks for processing text from high-dimensional primary care databases.

Recent developments in open-weight Small Language Models (SLMs), typically defined as models under 10 billion parameters, have emerged as a viable alternative, offering competent reasoning capabilities at reduced computational cost and enabling local execution on standard computing hardware [21]. This approach eliminates the transmission of PHI to third parties and substantially reduces economic costs.

Having the capacity to extract patient clinical features is particularly relevant for observational studies on acute infectious diseases, where key clinical indicators, including systemic fever, localised pain, and point-of-care dipstick parameters (nitrites, leukocytes, and haematuria), are rarely captured as discrete diagnostic codes in routine primary care.

To date, few studies have evaluated the real-world deployment of privacy-preserving local SLM pipelines for multilingual primary care EHR extraction, particularly in Southern European linguistic contexts. Therefore, the primary objective of this study was to develop, implement, and validate a privacy-preserving local pipeline to extract six uncoded clinical features: fever, nitrites, leukocytes, lumbar pain, abdominal pain, and haematuria, from primary care MEAP narratives in Catalonia [22].

## Methods

### Study Setting and Data Source

The study utilized clinical free-text records extracted from the Information System for the Development of Research in Primary Care (SIDIAP, Sistema d’Informació per al Desenvolupament de la Investigació en Atenció Primària) database in Catalonia [23].

The SIDIAP database is a primary care electronic health record system managed by the Institut d’Investigació en Atenció Primària Jordi Gol (IDIAPJGol) in Catalonia, Spain [24]. It contains pseudonymized information from different data sources: 1) EHR in Primary Health Care of the Catalan Health Institute (ICS: Institut Català de la Salut) (approximately attending 75% of the Catalan population); including sociodemographic characteristics, comorbidities registered as International Classification of Diseases, 10th revision, Clinical Modification (ICD­10-CM) codes [5], specialist referrals, clinical parameters, tobacco and alcohol consumption, sickness leave, date of death, laboratory test, and drug prescriptions issued in Primary Health Care, registered as Anatomical Therapeutic Chemical (ATC) classification system codes [6]. 2) Pharmacy invoice data corresponding to the Primary Care drug prescriptions, classified according to the ATC classification. 3) Pharmacy invoice data corresponding to the drug prescriptions dispensed in hospital pharmacies (MHDA database) [25].

In the context of the SIDIAP database, MEAP is the acronym that defines the semi-structured narrative format of free-text notes recorded by primary care physicians during patient visits [8]. MEAP stands for: M (Motiu de consulta – reason for encounter), E (Resultat de l’exploració – physical examination findings), A (Avaluació – assessment), and P (Planificació – plan). These narratives usually include the consultation reason, physical examination findings, point-of-care laboratory results, treatment prescriptions, and specific clinical symptoms, enabling access to rich patient information that is not captured through closed diagnostic codes, such as ICD-10-CM or ATC codes.

Based on their high prevalence and clinical relevance for pyelonephritis prediction [22], six key clincal features absent as structured codes were selected from MEAP records: fever, nitrites, leukocytes, lumbar pain, abdominal pain, and haematuria. This selection was established through a formal consensus panel of four experts: two primary care clinicians and two clinical pharmacologists.

The study population was a matched nested case-control selected from SIDIAP as part of the ITUCAT study, an observational study that included patients diagnosed with urinary tract infections (UTI) between 2012 and 2021 in Catalonia [26], [27], [28]. From this source population, we selected all patients aged 18 years or older attended at ICS primary care centres with at least one recorded UTI and a minimum follow-up of 45 days after the first UTI diagnosis, with matching based on age, year of the first UTI episode, and nursing home residency status. Cases were patients with an initial UTI diagnosis who subsequently met the study definition of acute pyelonephritis (APN) within 45 days, whereas controls were those who did not met APN definition within the same period. MEAP narratives were retrieved for all selected patients and used for feature extraction.

### MEAP Record Preprocessing

Raw MEAP records required automated cleaning to remove noise and non-relevant metadata, such as patient identification or dates, that could interfere with the language model inference. Preprocessing was implemented in Python using three sequential steps: line segmentation and delimiter parsing, whereby the free-text field was split across line breaks and pipe symbols (|) to isolate pure clinical text blocks from administrative tags (e.g., patient ID); temporal filtering, which restricted the text to a ±7-day window relative to the index diagnosis date (time zero), discarding entries outside this timeframe; and patient file disaggregation, in which the cleaned entries for each patient were separated into individual structured files to enable isolated per-patient processing.

### Privacy-Preserving Local Setup

To guarantee strict compliance with data protection laws and the ethical committee, sending clinical narratives of the patient to external servers was prohibited. Instead, we deployed a local inference environment using Ollama (Version 0.1.32, https://ollama.com) on a secure institutional server, the Catalan Health Institute (ICS) network.

The selected model was Phi4-mini, an open lightweight language model with approximately 3.8 billion parameters developed by Microsoft [29]. It was selected for its efficiency (suitable for local hardware with less than 12 GB VRAM), multilingual capability (pretrained on a diverse corpus including Catalan and Spanish, the predominant language in MEAPs), native function-calling/structured output capabilities and open-source availability. No data left the institutional firewall at any stage of processing.

### Features Extraction Pipeline and Prompt Engineering

The extraction workflow was implemented in Python. A structured output schema was defined for each target feature, consisting of a boolean variable indicating feature presence and a string variable for the corresponding narrative evidence. The local language model, Phi4-mini, accessed via Ollama, was configured with parameters governing token limits and response timeouts to optimise memory usage. All outputs were constrained to JSON format. The extraction was performed at the patient level: the full MEAP text for each patient was passed through the pipeline separately for each clinical feature, and this process was repeated iteratively until all features had been extracted. Patients without MEAP text were considered as not having any of the clinical feature. Figure 1 provides a schematic overview of the pipeline.

**Figure 1.**
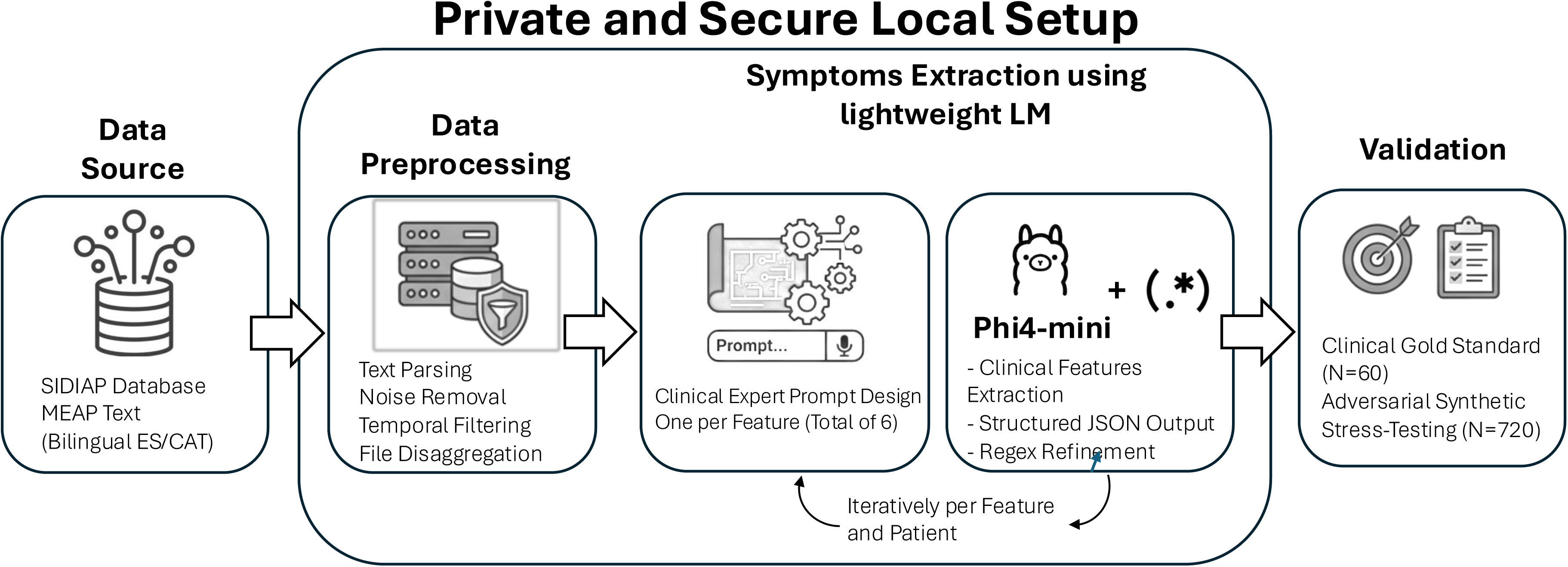
Extraction pipeline scheme

The prompting strategy was developed with four clinical experts (two primary care physicians and two clinical pharmacologists), with each clinical feature receiving a custom prompt that included explicit mentions, synonyms, regional terminology, and clearly specified inclusion and exclusion criteria. Under the guidance of an AI specialist, the prompt was refined and optimized using GPT-4.1 to maximize performance on lightweight local models like Phi4-mini. Over three iterative rounds, we used a test set of 20 MEAP narratives, which were excluded from the validation phase, to obtain the final prompt. For example, Figure 2 illustrated the prompt used for fever extraction (the full set of prompts is provided in Supplemental Table 1).

**Figure 2.**
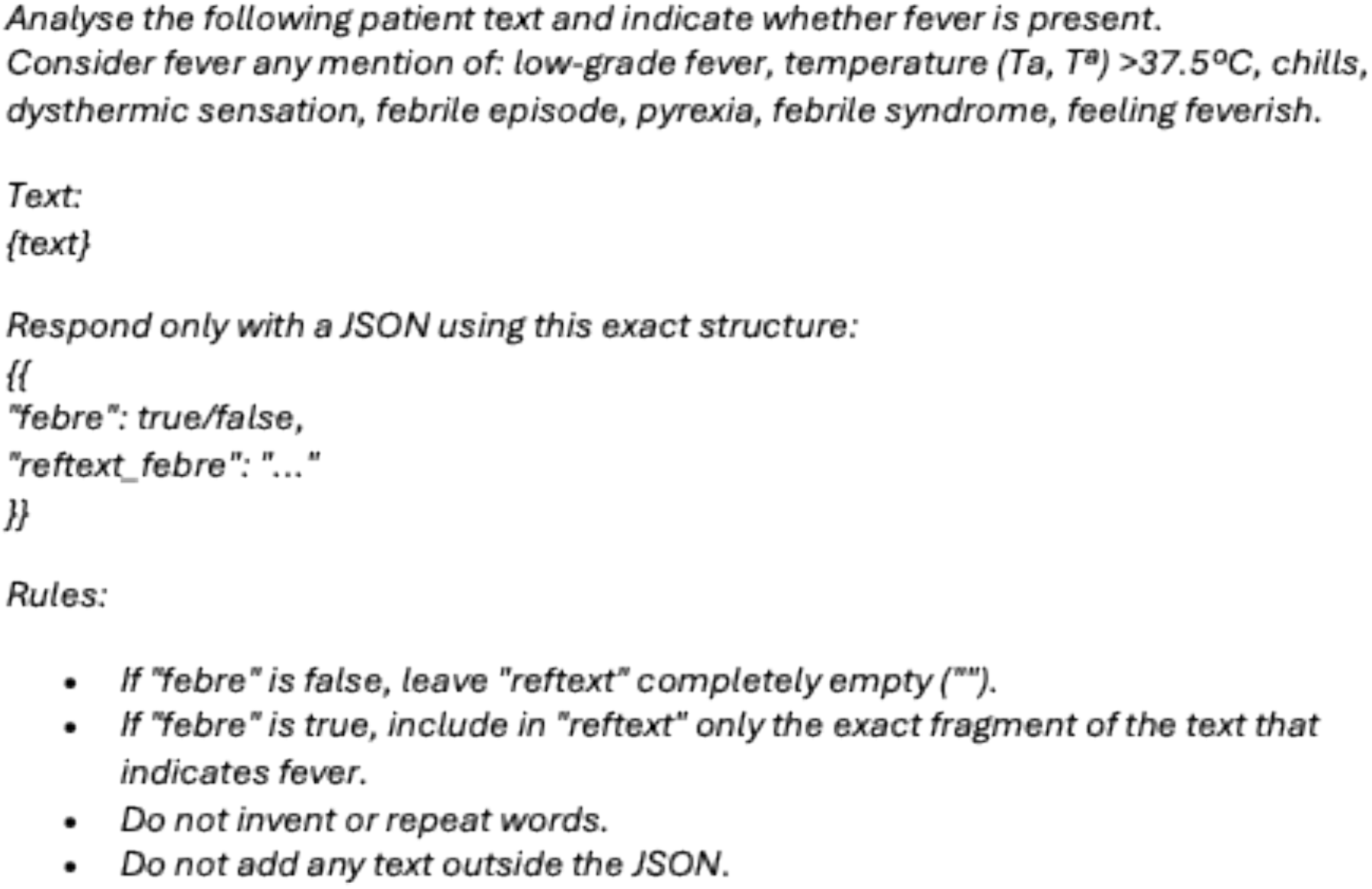
Prompt used for fever extraction

### Refinement Using Regular Expressions

Following the initial LLM inference, we identified systematic error patterns, particularly in handling edge cases, clinical shorthand, and quantitative laboratory parameters. To enhance pipeline performance, we implemented a post-processing module that combined LLM outputs with deterministic rule-based logic using regular expressions. The rules were developed manually and iteratively based on observed error patterns and applied consistently across all features. In cases of conflict, the regular expression output was prioritised (Supplemental Table 1).

All prompts and regular expressions used in the pipeline are provided in the Supplementary Material.

### Validation Strategy

The pipeline was validated across two independent studies. The first was a real-world clinical gold standard validation using a stratified random sample of 60 real patients, 30 cases of confirmed pyelonephritis and 30 controls with urinary tract infection, with its MEAP records. Each group included 10 patients presenting with more than two clinical features, 10 with a single clinical feature, and 10 without any clinical features. Four primary care clinicians performed blinded manual reviews, and each record was independently evaluated by a pair of reviewers to assess inter-reviewer agreement. The gold standard label was defined as the consensus of the two reviewers. In case of disagreement, a new manual revision was performed together with all four experts only for fever and lumbar pain, and the rest were discarded from the validation study. To assess whether Phi4-mini performs similarly to alternative small language models, an additional analysis was conducted using a second open-weight SLM (Gemma3:4b [30]) on the same 60 clinical records from the gold standard validation.

The second was an adversarial synthetic stress-testing validation using a dataset of 720 clinical-style texts. For each of the target clinical features, 60 texts were generated using commercial large language models: GPT 4.1 turbo (OpenAI. (2026). ChatGPT (GPT-4.1 version) [Large language model]. https://chatgpt.com) and Grok 4.1 (xAI. (2026). Grok (Grok 4.1 version) [Large language model]. https://grok.com) [31]. The synthetic narratives were enriched in features to achieve a prevalence different from that observed in the real texts and to provide more data for testing pipeline performance. We therefore instructed the LLMs to generate texts with an overall prevalence close to 50%. Texts were designed to include rare synonyms, syntactic variations, code-switching and adversarial examples (negation traps, ambiguous references, overlapping features) to specifically challenge lightweight models such as Phi4-mini (the prompts are available in Supplemental Table 3). This set was processed through the pipeline without any retraining or prompt adjustment. The extraction pipeline was evaluated against the ground-truth labels provided during synthetic generation.

### Statistical Methods

The frequency of detected clinical features was summarised using absolute and relative frequencies. The distribution and co-occurrence patterns were explored using UpSet plots, with separate visualisations generated for cases and controls. Differences between cases and controls in feature distribution were assessed using standardised mean differences (SMD). The feature profiles at a patient level and between study groups were also used as a qualitative validation of the extraction pipeline’s performance.

To assess performance in the validation phase, confusion matrices were used to estimate standard metrics, including prevalences, sensitivity, specificity, accuracy, positive predictive value, and negative predictive value (along with its 95% confidence intervals (CI) using exact methods [32]). Inter-reviewer agreement, between manual reviewers and between SLMs, was evaluated using Cohen’s kappa [33].

### Ethical Considerations

The study was conducted in accordance with the Declaration of Helsinki, Good Research Practice principles and guidelines, and the Real Decreto 957/2020, of November 3, which regulates observational studies of medicines for human use. This study was approved by the Ethics Committee for Research with Medicines (CEIm) of the Jordi Gol i Gurina Foundation (IDIAPJGol), with ethical approval code 24/196, protocol code IJG-ITUCAT-2022 (WP3), on 24 July 2025. An amendment was approved on 26 November 2026. All data was stored, processed and analysed in our secure institutional server.

## Results

### Study Population with its MEAP Narratives

The final study population comprised 1,481 matched case-control pairs (2,962 patients), of whom 486 were male and 995 were female. A total of 15,498 MEAP narratives were available for the selected patients and used for extraction.

### Features Extracted Distribution

Table 1 shows the distribution of features detected by the pipeline, overall and by case/control status. A total of 4,663 clinical features were detected, corresponding to 1,883 patients (63.6% of the total population). The most prevalent features were leukocytes (43.5%) and haematuria (40.4%), followed by fever (21.0%). The distribution was imbalanced between cases and controls, with cases having a higher proportion of features than controls (71.3% vs. 55.8%; SMD = 0.326). This pattern was also observed at the feature level, particularly for fever (488 cases [33.0%] vs. 133 controls [9.0%]; SMD = 0.616) and lumbar pain (430 cases [29.0%] vs. 145 controls [9.8%]; SMD = 0.502).

**Table 1.** Distribution of the extracted clinical features.

|  | Overall | Cases | Controls | SMD* |
| --- | --- | --- | --- | --- |
| **N** | 2962 | 1481 | 1481 |  |
| **Presence of clinical feature, n (%)** | 1883 (63.6) | 1056 (71.3) | 827 (55.8) | 0.326 |
| **Clinical feature, n (%)** |  |  |  |  |
| **Fever** | 621 (21.0) | 488 (33.0) | 133 ( 9.0) | 0.616 |
| **Nitrites** | 505 (17.0) | 323 (21.8) | 182 (12.3) | 0.255 |
| **Leukocytes** | 1288 (43.5) | 709 (47.9) | 579 (39.1) | 0.178 |
| **Lumbar Pain** | 575 (19.4) | 430 (29.0) | 145 ( 9.8) | 0.502 |
| **Abdominal Pain** | 476 (16.1) | 309 (20.9) | 167 (11.3) | 0.263 |
| **Haematuria** | 1198 (40.4) | 693 (46.8) | 505 (34.1) | 0.261 |

\* SMD>0.1 considered indicative of imbalance between groups

The UpSet plot (Figure 3) shows the distribution of combined clinical features at the patient level. Overall, the most frequent combination was haematuria with leukocytes (268 patients, 9.0% of the total population), followed by the triple combination of nitrites, haematuria, and leukocytes (116 patients, 3.9%). Single features were also observed, with haematuria, leukocytes, and fever being the most prevalent (150 (5.1%), 140 (4.7%), and 91 (3.1%), respectively). Feature profiles differed between cases and controls (Supplemental Figures 1 and 2). Cases exhibited more complex profiles, with a higher proportion of patients presenting with more than one feature, whereas in controls, patients with a single-feature were more prevalent.

**Figure 3.**
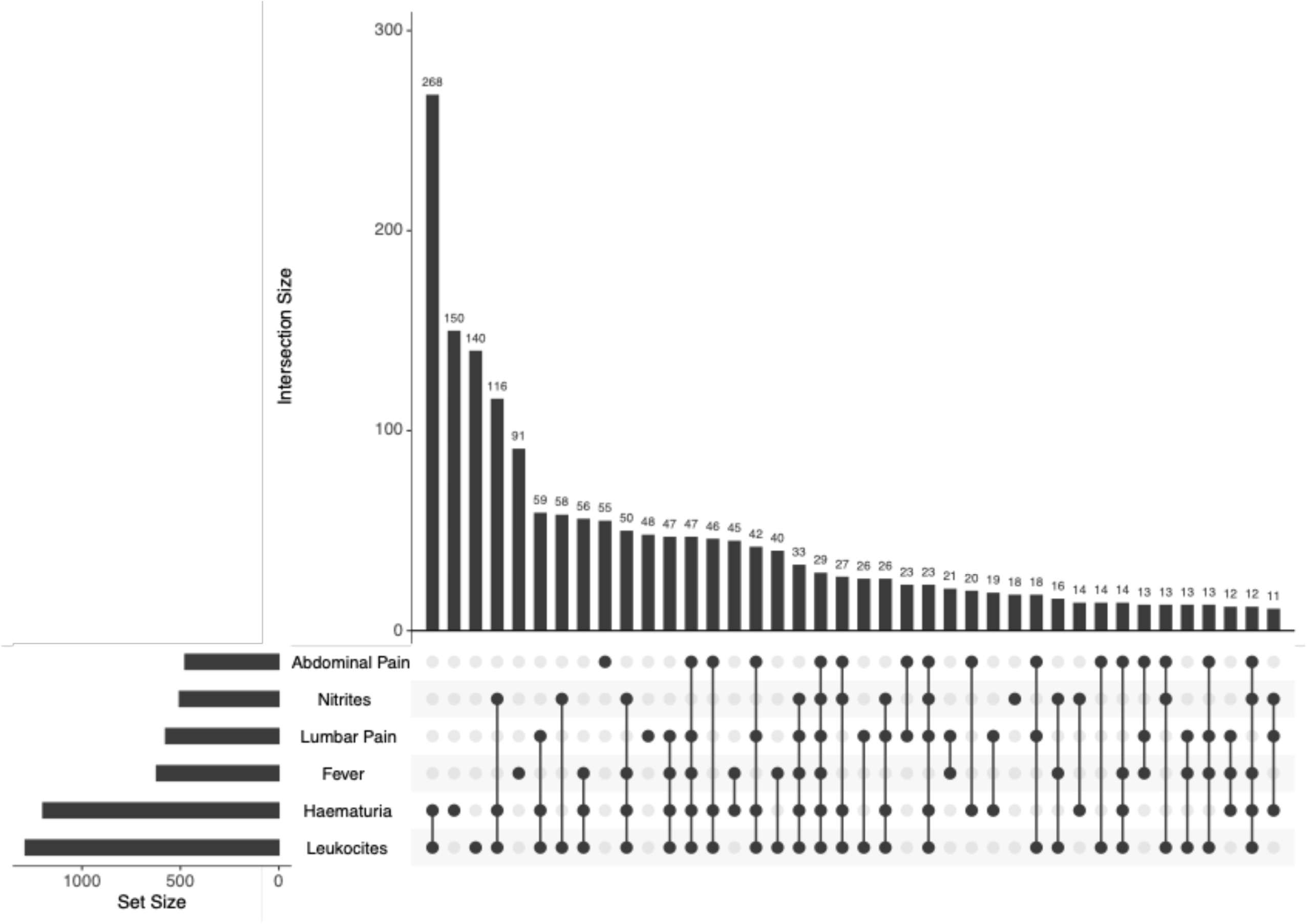
Upset chart of extracted clinical features

### Real-World Validation Performance

The performance of the pipeline was evaluated against the double-blind clinician gold standard for the six target clinical features. Inter-reviewer agreement was 84.4% (Cohen’s kappa). After re-evaluation of discordant pairs, 23 narratives were excluded. The remaining MEAP narratives yielded 73 gold-standard features (prevalence: 21.7%; Table 2).

**Table 2.** Performance metrics from gold standard EHR MEAP narratives.

|  | TP | TN | FP | FN | Apparent Prevalence | True Prevalence | Sensitivity | Specificity | Accuracy | PPV | NPV |
| --- | --- | --- | --- | --- | --- | --- | --- | --- | --- | --- | --- |
| **Global** | 61 | 255 | 9 | 12 | 20.8 [ 16.6 , 25.5 ] | 21.7 [ 17.4 , 26.4 ] | 83.6 [ 73.0 , 91.2 ] | 96.6 [ 93.6 , 98.4 ] | 93.8 [ 90.6 , 96.1 ] | 87.1 [ 77.0 , 93.9 ] | 95.5 [ 92.3 , 97.7 ] |
| **Clinical feature:** |  |  |  |  |  |  |  |  |  |  |  |
| **Fever** | 5 | 51 | 0 | 4 | 8.3 [ 2.8 , 18.4 ] | 15.0 [ 7.1 , 26.6 ] | 55.6 [ 21.2 , 86.3 ] | 100.0 [ 93.0 , 100.0 ] | 93.3 [ 83.8 , 98.2 ] | 100.0 [ 47.8 , 100.0 ] | 92.7 [ 82.4 , 98 ] |
| **Nitrites** | 7 | 43 | 1 | 0 | 15.7 [ 7.0 , 28.6 ] | 13.7 [ 5.7 , 26.3 ] | 100.0 [ 59.0 , 100.0 ] | 97.7 [ 88.0 , 99.9 ] | 98.0 [ 89.6 , 100.0 ] | 87.5 [ 47.3 , 99.7 ] | 100.0 [ 91.8 , 100.0 ] |
| **Leukocytes** | 14 | 33 | 1 | 3 | 29.4 [ 17.5 , 43.8 ] | 33.3 [ 20.8 , 47.9 ] | 82.4 [ 56.6 , 96.2 ] | 97.1 [ 84.7 , 99.9 ] | 92.2 [ 81.1 , 97.8 ] | 93.3 [ 68.1 , 99.8 ] | 91.7 [ 77.5 , 98.2 ] |
| **Lumbar Pain** | 7 | 50 | 0 | 3 | 11.7 [ 4.8 , 22.6 ] | 16.7 [ 8.3 , 28.5 ] | 70.0 [ 34.8 , 93.3 ] | 100.0 [ 92.9 , 100.0 ] | 95.0 [ 86.1 , 99.0 ] | 100.0 [ 59.0 , 100.0 ] | 94.3 [ 84.3 , 98.8 ] |
| **Abdominal Pain** | 8 | 47 | 0 | 2 | 14.0 [ 6.3 , 25.8 ] | 17.5 [ 8.7 , 29.9 ] | 80.0 [ 44.4 , 97.5 ] | 100.0 [ 92.5 , 100.0 ] | 96.5 [ 87.9 , 99.6 ] | 100.0 [ 63.1 , 100.0 ] | 95.9 [ 86.0 , 99.5 ] |
| **Haematuria** | 20 | 31 | 7 | 0 | 46.6 [ 33.3 , 60.1 ] | 34.5 [ 22.5 , 48.1 ] | 100.0 [ 83.2 , 100.0 ] | 81.6 [ 65.7 , 92.3 ] | 87.9 [ 76.7 , 95.0 ] | 74.1 [ 53.7 , 88.9 ] | 100.0 [ 88.8 , 100.0 ] |

TP: True Positive, TN: True Negative, FP: False Positive, FN: False Negative, PPV/NPV: Positive/Negative Predictive Value

Overall performance (95% CI) was as follows: accuracy 93.8% (90.6–96.1), with 9 false positives and 12 false negatives; sensitivity 83.6% (73.0–91.2); specificity 96.6% (93.6–98.4); PPV 87.1% (77.0–93.9); and NPV 95.5% (92.3–97.7). Nitrites, abdominal pain, and lumbar pain achieved the highest accuracy (98.0%, 96.5%, and 95.0%, respectively), while haematuria showed the lowest accuracy (87.9% [76.7–95.0]), mainly due to seven false positives (PPV: 74.1% [53.7–88.9]). Performance was consistent between cases and controls (Supplemental Table 2).

Agreement between the two SLMs was moderate to high, with an overall Cohen’s kappa of 66.9 (47.4–86.4), although it varied across features. Fever showed the lowest agreement (35.0 [11.2– 58.8]), whereas lumbar pain and leukocytes showed the highest agreement (Supplemental Table 4). The performance metrics of the alternative SLM (Gemma 3:4b) on the 60 MEAP narratives are presented in Supplemental Table 5. In overall, observed accuracy was 84.9% (80.6–88.5), sensitivity was 97.3% (90.5–99.7) with 2 false negatives, and specificity was 81.4% (76.2–85.9) with 49 false positives.

### Adversarial Synthetic Evaluation

The stress-test was conducted on 720 synthetic notes (360 generated by GPT-4.1 and 360 by Grok-4.1). As expected, the prevalence was close to 50% (47.9%), with a total of 345 features detected. Fever, abdominal pain, and haematuria reached the target prevalence, whereas leukocytes were the least prevalent (42.5%; Table 3).

**Table 3.** Performance metrics from adversarial synthetic validation.

|  | TP | TN | FP | FN | Apparent<br>Prevalence | True<br>Prevalence | Sensitivity | Specificity | Accuracy | PPV | NPV |
| --- | --- | --- | --- | --- | --- | --- | --- | --- | --- | --- | --- |
| **Global** | 255 | 373 | 2 | 90 | 35.7 [ 32.2 ,<br>39.3 ] | 47.9 [ 44.2 ,<br>51.6 ] | 73.9 [ 68.9 , 78.5 ] | 99.5 [ 98.1 , 99.9 ] | 87.2<br>[ 84.6 , 89.6 ] | 99.2<br>[ 97.2 , 99.9 ] | 80.6<br>[ 76.7 , 84.1 ] |
| **Clinical feature:** |  |  |  |  |  |  |  |  |  |  |  |
| **Fever** | 26 | 60 | 0 | 34 | 21.7 [ 14.7 ,<br>30.1 ] | 50.0 [ 40.7 ,<br>59.3 ] | 43.3 [ 30.6 , 56.8 ] | 100.0 [ 94.0 , 100.0<br>] | 71.7 [ 62.7 , 79.5 ] | 100.0 [ 86.8 ,<br>100.0 ] | 63.8 [ 53.3 ,<br>73.5 ] |
| **Nitrites** | 44 | 64 | 1 | 11 | 37.5 [ 28.8 ,<br>46.8 ] | 45.8 [ 36.7 ,<br>55.2 ] | 80.0 [ 67.0 , 89.6 ] | 98.5 [ 91.7 , 100.0 ] | 90.0 [ 83.2 , 94.7 ] | 97.8 [ 88.2 ,<br>99.9 ] | 85.3 [ 75.3 ,<br>92.4 ] |
| **Leukocytes** | 42 | 68 | 1 | 9 | 35.8 [ 27.3 ,<br>45.1 ] | 42.5 [ 33.5 ,<br>51.9 ] | 82.4 [ 69.1 , 91.6 ] | 98.6 [ 92.2 , 100.0 ] | 91.7 [ 85.2 , 95.9 ] | 97.7 [ 87.7 ,<br>99.9 ] | 88.3 [ 79.0 ,<br>94.5 ] |
| **Lumbar Pain** | 53 | 61 | 0 | 6 | 44.2 [ 35.1 ,<br>53.5 ] | 49.2 [ 39.9 ,<br>58.4 ] | 89.8 [ 79.2 , 96.2 ] | 100.0 [ 94.1 , 100.0<br>] | 95.0 [ 89.4 , 98.1 ] | 100.0 [ 93.3 ,<br>100.0 ] | 91.0 [ 81.5 ,<br>96.6 ] |
| **Abdominal Pain** | 35 | 60 | 0 | 25 | 29.2 [ 21.2 ,<br>38.2 ] | 50.0 [ 40.7 ,<br>59.3 ] | 58.3 [ 44.9 , 70.9 ] | 100.0 [ 94.0 , 100.0<br>] | 79.2 [ 70.8 , 86.0 ] | 100.0 [ 90.0 ,<br>100.0 ] | 70.6 [ 59.7 ,<br>80.0 ] |
| **Haematuria** | 55 | 60 | 0 | 5 | 45.8 [ 36.7 ,<br>55.2 ] | 50.0 [ 40.7 ,<br>59.3 ] | 91.7 [ 81.6 , 97.2 ] | 100.0 [ 94.0 , 100.0<br>] | 95.8 [ 90.5 , 98.6 ] | 100.0 [ 93.5 ,<br>100.0 ] | 92.3 [ 83.0 ,<br>97.5 ] |

TP: True Positive, TN: True Negative, FP: False Positive, FN: False Negative, PPV/NPV: Positive/Negative Predictive Value Table 3 Performance metrics from adversarial synthetic validation

Overall accuracy was 87.2% (95% CI: 84.6–89.6), with 2 false positives and 90 false negatives, corresponding to a specificity of 99.5% (95% CI: 98.1–99.9), PPV of 99.2% (95% CI: 97.2–99.9), NPV of 80.6% (95% CI: 76.7–84.1), and sensitivity of 73.9% (95% CI: 68.9–78.5). Performance varied at the feature level: most features achieved an accuracy above 90%, except for fever (71.7% [95% CI: 62.7–79.5]) and abdominal pain (79.2% [95% CI: 70.8–86.0]), which showed the lowest performance. Results were nearly identical across the two language models (data not shown).

## Discussion G Conclusions

### Principal Findings

Across 15,498 MEAP records from a matched cohort of 2,962 patients, the pipeline identified 4,663 individual clinical feature occurrences, corresponding to 63.6% of the study population. The extracted feature distribution demonstrated strong clinical face validity, showing a significantly higher clinical feature burden in cases compared to non-progressing controls (71.3% vs. 55.8%; SMD = 0.326), particularly for classic systemic signs such as fever (SMD = 0.616) and lumbar pain (SMD = 0.502).

In real-world clinical evaluation against a double-blind clinician gold standard, the pipeline demonstrated high overall accuracy (93.8%), specificity (96.6%), and negative predictive value (95.5%). Performance was especially robust for laboratory-adjacent parameters and localized pain indicators, including nitrites (98.0% accuracy), abdominal pain (96.5% accuracy), and lumbar pain (95.0% accuracy). When subjected to adversarial synthetic stress-testing across 720 challenging narratives enriched with syntactic noise, regional dialects, and negation traps, the pipeline maintained near-perfect specificity (99.5%) and positive predictive value (99.2%), confirming its resilience against false-positive hallucinations.

### Comparison with Prior Work

Proprietary cloud models such as GPT-4 and ChatGPT-3.5 have established strong benchmarks in parsing clinical free-text narratives [34], [35], [36], [37], achieving, for example, 89% overall accuracy in lung cancer classification and high concordance (>90% accuracy) in glioblastoma and colorectal cancer datasets [37]. However, transmitting patient-level Electronic Health Records (EHRs) to external commercial APIs introduces substantial regulatory barriers and privacy risks. In response, recent efforts have prioritised open-source and open-weight architectures for local, privacy-preserving extraction [35], [38], [39]. Notably, Bai et al. [40] evaluated open-weight models using a multimodule framework with post-processing for cardiorespiratory clinical feature extraction. In their evaluation on clinical notes containing cardiorespiratory features (n=69), gpt-oss-120B and Llama 3.3-70B achieved precision of 0.97 and 0.89, respectively. Another study using hybrid RAG–GPT-4 pipelines on MIMIC-III achieved substance use mention identification with up to 0.99 precision [41]. Similarly, locally executed Llama 3.3-70B with prompt engineering yielded high performance for genitourinary symptom extraction (F₁: 0.92; precision: 0.89; recall: 0.96). However, under fully zero-shot local conditions, the same model showed lower performance for clinical feature extraction (F₁: 0.78; precision: 0.71; recall: 0.87) [42], [43].

Domain-adapted small language models (SLMs) increasingly rival large foundation models when explicitly infused with structured clinical knowledge and domain-specific reasoning [21]. Kim et al. [44] demonstrated that training SLMs on high-quality medical textbook data and synthetic chain-of-thought reasoning paths dramatically elevates clinical performance; their model, Meerkat-7B, performed similarly to other LLMs and even surpassed GPT-3.5. Similarly, Corso et al. [45] established that combining clinician expertise with targeted prompt engineering is the decisive factor in enabling SLMs to accurately extract cancer information from unstructured EHRs. Furthermore, several studies suggest that combining probabilistic LLMs with deterministic, rule-based regular expressions can optimise extraction fidelity by mitigating hallucinations and capturing standardised clinical terminology [46], [47].

Our findings align directly with this line of work, demonstrating that a compact 3.8-billion-parameter SLM (Phi4-mini) executed locally on modest hardware, can achieve high extraction accuracy for complex clinical variables, comparable to that of larger architectures.

### Strengths and Limitations

Our approach leverages lightweight open models optimised for resource-constrained hardware. Combining probabilistic SLM reasoning with deterministic regex post-processing mitigates hallucination errors and effectively captures domain-specific clinical terminology in bilingual environments (Catalan/Spanish).

A major strength of this study is its strict privacy-preserving architecture. By deploying the open-weight model via Ollama within the Catalan Health Institute (ICS) network, zero patient health information crossed institutional boundaries, ensuring full local regulatory compliance while eliminating recurring per-token fees. Another strength is the combined expertise of AI specialists and clinicians, which enabled the development of prompt engineering specifically curated for the narrative style of primary care notes, likely enhancing extraction accuracy. Methodologically, the dual-validation strategy provides both clinical fidelity and a rigorous assessment of edge-case failure modes.

Furthermore, while most existing clinical NLP tools are tailored exclusively for English-language EHRs, our pipeline effectively processes multilingual clinical text combining Catalan and Spanish, accommodating the code-switching and informal clinical shorthand typical of primary care notes in Catalonia. This approach offers a practical, privacy-compliant solution for extracting granular clinical data from primary care EHRs, which are often underutilised in research due to the challenges of unstructured data.

Several clinical and methodological limitations should be considered. First, as a 3.8-billion-parameter model, Phi4-mini exhibits lower reasoning capacity than massive frontier LLMs, making it more vulnerable to narrative noise, non-standard clinical shorthand, complex negations, and batch throughput bottlenecks during large-scale local execution. This trade-off was observed when comparing Phi4-mini with Gemma 3:4b, although agreement between models was high, Gemma 3:4b showed higher sensitivity at the cost of a higher number of false positives. This performance variability across SLMs of similar size suggests that model selection is a critical design decision. Second, prioritisation of regex outputs over LLM predictions in cases of conflict maximised specificity (preventing false attribution) but likely contributed to conservative feature capture. Third, during the real-world clinician validation phase, 23 narrative evaluations (6.4% of the validation sample) were excluded due to persistent inter-annotator disagreement between the two blinded clinical reviewers. While discarding unresolved ambiguous cases ensured a robust and definitive gold standard for pipeline evaluation, this approach inherently introduces a degree of selection bias that should be acknowledged. Fourth, the six extracted variables represent a mixture of symptoms, clinical signs, and urinalysis findings; therefore, they should not be interpreted as equivalent clinical manifestations or as independent diagnostic criteria for pyelonephritis. Fever and lumbar/flank pain are clinically relevant features of systemic UTI, whereas nitrite and leukocyte positivity provide urinalysis evidence of urinary inflammation or bacteriuria and do not, in isolation, distinguish cystitis from pyelonephritis. Fifth, the lower sensitivity observed for fever and abdominal pain may reflect both model limitations and variability in clinical documentation. Fever may be recorded as a numerical temperature, as an indirect description, or using terminology such as afebrile, while abdominal pain may be documented using heterogeneous anatomical descriptions. In addition, the relatively lower specificity for haematuria may reflect ambiguity in documentation, including patient-reported bleeding, dipstick blood, microscopic haematuria, historical findings, or bleeding not attributable to UTI. Sixth, MEAP notes are highly subjective and clinician-dependent; not all symptoms and signs are recorded, and in many cases, no clinical features are documented at all. The completeness of MEAP documentation is highly heterogeneous and depends on the individual clinician, the time available per consultation, and the overall volume of visits.

Finally, although the pipeline was validated on a large multi-centre primary care cohort (SIDIAP), the evaluated notes were restricted to a specific clinical context (urinary tract infections). Future work should evaluate the generalisability of this local hybrid approach across broader clinical domains, such as cardiovascular and respiratory conditions, and consider periodic re-evaluation as clinical documentation practices evolve.

### Clinical and Methodological Implications

Structured codes capture only a fraction of clinical reality; the remaining narrative content is often discarded or manually abstracted at high cost. From a clinical perspective, this work provides a reproducible pipeline for healthcare institutions seeking to extract uncoded variables from free-text records, offering researchers access to a substantial volume of previously underutilised data in primary care EHR research. The ability to systematically capture these features enables the enrichment of longitudinal patient profiles with uncoded clinical information, which is particularly relevant for acute infections, where timely and complete data can inform clinical decision-making. Predictions based on structured information, such as comorbidities and patient characteristics, may be insufficient, and the incorporation of clinical features represents a substantial improvement, as these undoubtedly influence the physician’s assessment of patient risk and, consequently, their therapeutic decisions. In this study, we incorporated the six extracted clinical features: fever, nitrites, leukocytes, lumbar pain, abdominal pain, and haematuria; to enrich predictive risk models for severe complications, specifically acute pyelonephritis, within the ITUCAT study [27].

In addition, the potential utility of this approach extends other types of uncoded information. It could also be applied to establish severity criteria or disease staging, as well as to detect adverse effects or to identify the reasons behind changes in treatment decisions.

Our pipeline offers a transferable template for other healthcare systems (for example, hospital frameworks) facing similar constraints. It is modular and configurable, meaning that prompts and regular expressions can be adapted to new clinical domains or languages. Furthermore, its applicability extends beyond research: it could be used to support for routine management tasks, such as clinical documentation quality monitoring, patient profiling, or resource planning.

## Conclusions

This study demonstrates that a fully local, privacy-preserving hybrid pipeline, combining a small open-weight small language model, 3.8B parameter Phi4-mini, with deterministic regular expressions, provides an accurate, scalable, and fully compliant solution for extracting uncoded clinical features from multilingual primary care narratives. By achieving high overall accuracy and specificity without transmitting data outside institutional firewalls, the pipeline eliminates data privacy risks and offers a practical pathway for leveraging unstructured EHR data in real-world clinical research.

## Data Availability

The data produced in the present study are not available due to institutional and regulatory restrictions on the sharing of patient‑level clinical data.

