## Supplemental Figure 1 for "Uncoded Clinical Features from Multilingual Electronic Health Records in Catalonia: Development and Validation Study"

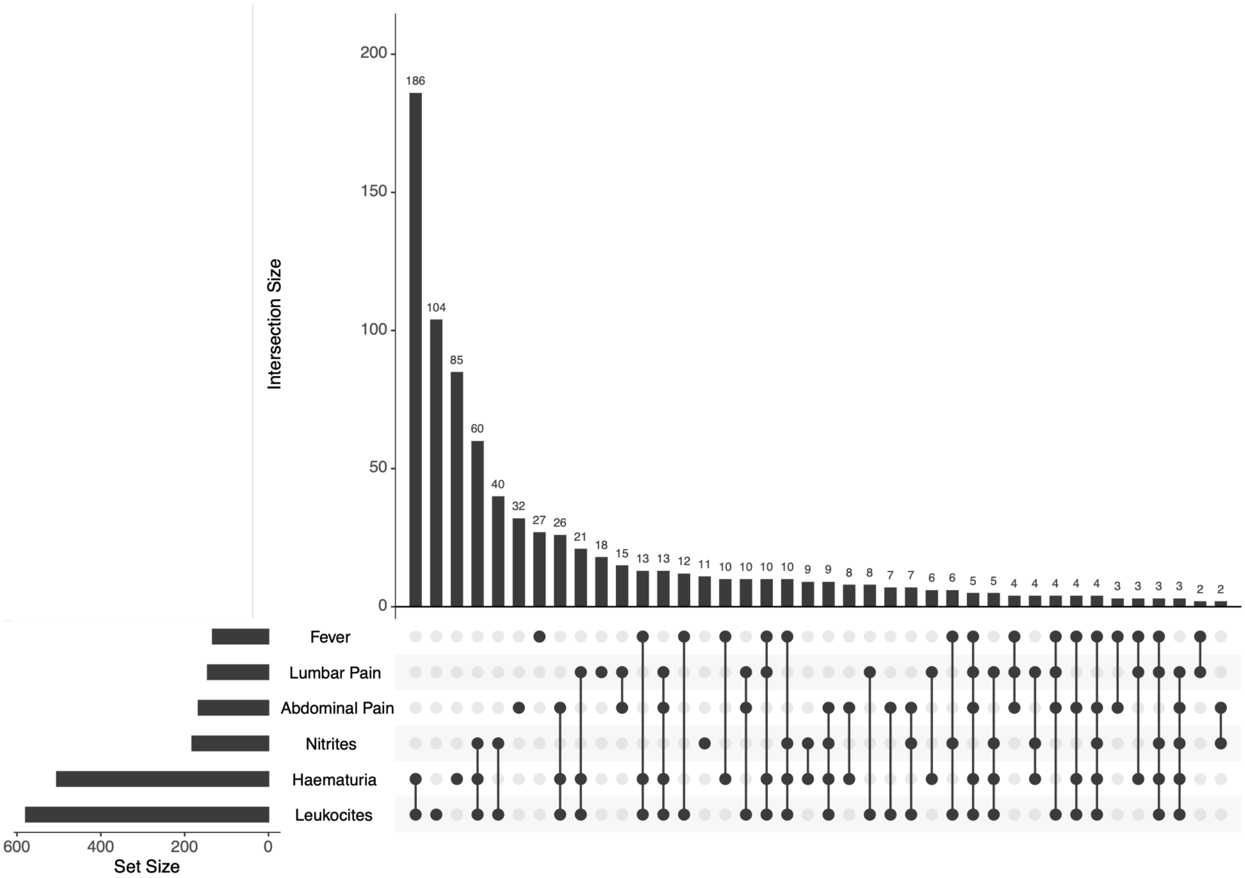


*Supplemental Figure 1 Clinical Features Distribution in Controls*

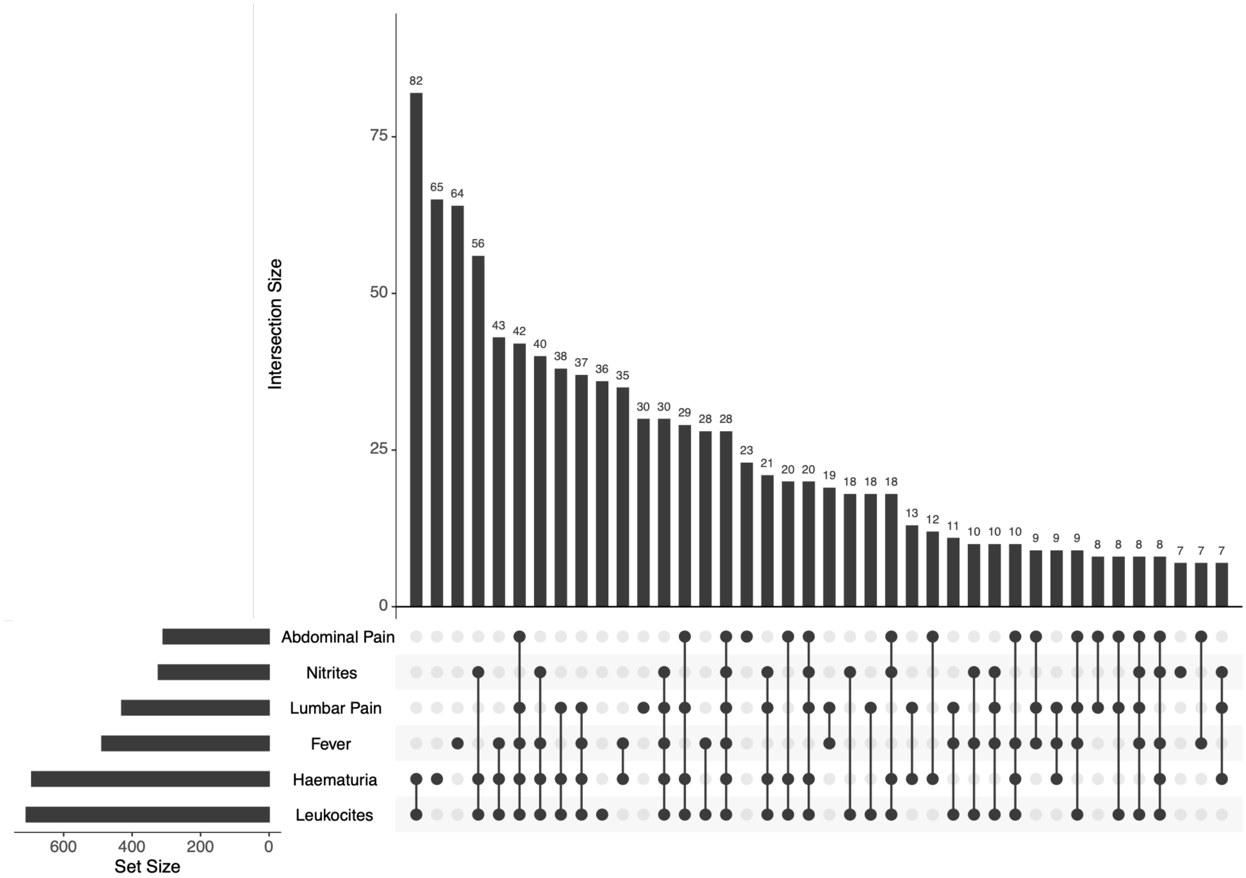


*Supplemental Figure 2. Clinical Features Distribution in Cases*

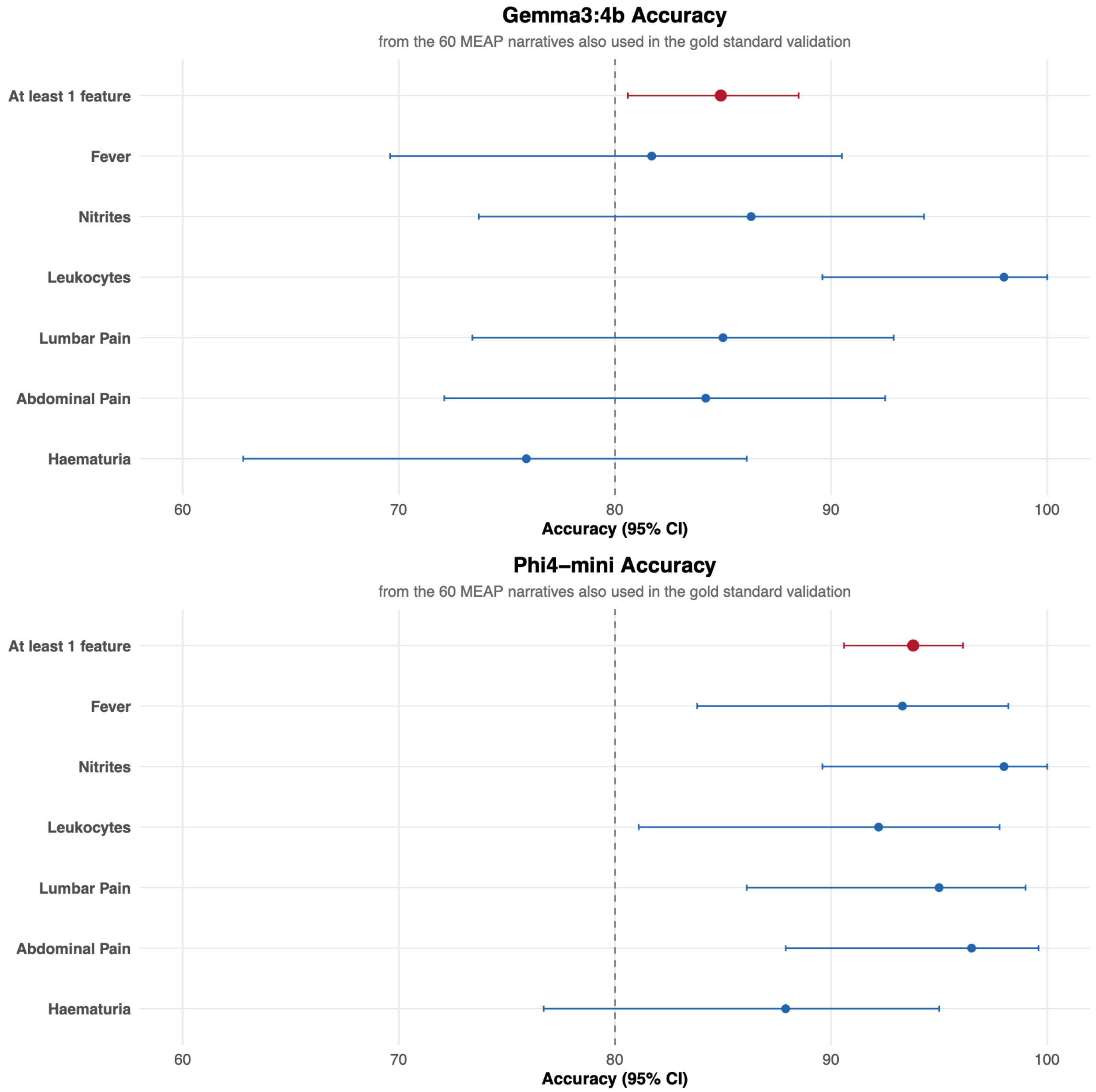


*Supplemental Figure 3 Accuracy in Gemma 3:4b and Phi4-mini*

### Supplementary Table 1. Clinical features-specific prompts and regular expressions used for extraction and refinement

<table>
<thead>
<tr><th>Clinical Feature</th><th>Keywords Provided by the Clinicians</th><th>Prompt Translated to English</th><th>Original Prompt</th><th>Regular Expression</th></tr>
</thead>
<tbody>
<tr>
<td>Fever</td>
<td>febrícula, temperatura (Ta, Tª) >37,5ºC, calfreds, sensació distèrmica, quadre febril, pirexia, sindrom febril, destemperat</td>
<td>Analyse the following patient text and indicate whether fever is present.<br>Consider fever any mention of: low-grade fever, temperature (Ta, Tª) >37.5ºC, chills, dysthermic sensation, febrile episode, pyrexia, febrile syndrome, feeling feverish.<br><br>Text:<br>{t}<br>Respond only with a JSON using this exact structure:<br>{{<br>"febre": true/false,<br>"reftext_febre": "..."<br>}}<br><br>Rules:<br>-If "febre" is false, leave "reftext" completely empty ("").<br>-If "febre" is true, include in "reftext" only the exact fragment of the text that indicates fever.<br>-Do not invent or repeat words.<br>-Do not add any text outside the JSON.</td>
<td>Analitza el següent text de pacient i indica si hi ha febre.<br>Considera febre qualsevol menció de: febrícula, temperatura (Ta, Tª) >37,5ºC, calfreds, sensació distèrmica, quadre febril, pirexia, sindrom febril, destemperat.<br><br>Text:<br>{t}<br>Respon només amb un JSON amb aquesta estructura exacta:<br>{{<br>"febre": true/false,<br>"reftext_febre": "..."<br>}}<br>Regles:<br>- Si "febre" és false, deixa "reftext" completament buit ("").<br>- Si "febre" és true, posa a "reftext" **només el fragment exacte del text** que indica febre.<br>- No inventis ni repeteixis paraules.<br>- No afegeixis cap text fora del JSON.</td>
<td>febre|febricula|temperatura\s*:?\s*>?\s*37\.?5|tª\s*:?\s*>?\s*37\.?5|ta\s*:?\s*>?\s*37\.?5|calfreds|sensaci[oó] dist[ée]rmica|quadre febril|pirexia|s[ií]ndrom febril|destemperat|fiebre|escalofr[ií]os|temperatura\s*:?\s*>?\s*37\.?5</td>
</tr>
</tbody>
</table>

<table>
<tr>
<td>Nitrites</td>
<td>Nitrites +</td>
<td>Analyse the following patient text and indicate whether nitrites are positive or negative.<br>Only consider them positive if the word "nitrits" (or "nitrits:") appears in the same fragment as "Positiu", "Positiva", or the symbol "+".<br>Only consider them negative if "Negatiu", "Negativa", or the symbol "-" appears in the same fragment as the word "nitrits".<br><br>Do not consider any "+" or "-" that is not directly related to nitrites.<br><br>Text:<br>{t}<br>Respond only with a JSON using this exact structure:<br>{{<br>"nitrits": true/false,<br>"reftext_nitrits": "..."<br>}}<br><br>Rules:<br>- If "nitrits" is false, leave "reftext" completely empty ("").<br>- If "nitrits" is true, include only the exact fragment of the text where positive nitrites appear.<br>- Do not invent or add words.<br>- Do not add any text outside the JSON.</td>
<td>Analitza el següent text de pacient i indica si els nitrits són positius o negatius.<br>Només considera que són positius si apareix la paraula "nitrits" (o "nitrits:" ) al mateix fragment que: "Positiu", "Positiva" o el símbol "+".<br>Només considera negatius si apareix "Negatiu", "Negativa" o el símbol "-" al mateix fragment que la paraula "nitrits".<br><br>No consideris cap "+" o "-" que no estigui relacionat directament amb nitrits.<br><br>Text:<br>{t}<br><br>Respon només amb un JSON amb aquesta estructura exacta:<br>{{<br>"nitrits": true/false,<br>"reftext_nitrits": "..."<br>}}<br><br>Regles:<br>- Si "nitrits" és false, deixa "reftext" completament buit ("").<br>- Si "nitrits" és true, posa només el fragment exacte del text on apareixen els nitrits positius.<br>- No inventis ni afegeixis paraules.<br>- No afegeixis cap text fora del JSON.</td>
<td>nitrits?\s*:?\s*\+\s*|nitrits?\s*:?\s*positiu|nitrits?\s*:?\s*positiva|nitrits?\s*:?\s*−|nitrits?\s*:?\s*negatiu|nitrits?\s*:?\s*negativa|nitritos?\s*:?\s*\+\s*|nitritos?\s*:?\s*positivo|nitritos?\s*:?\s*negativo</td>
</tr>
</table>

<table>
<tr>
<td>Leukocytes</td>
<td>Leukocytes and Traçes, +, ++ o +++.</td>
<td>Analyse the following patient text and indicate whether leukocytes are positive or negative.<br>Only consider them positive if the word "leucòcits" (or "leucocits:") appears in the same fragment as "Traçes", "+", "++", or "+++".<br>Only consider them negative if "Negatiu" or "Negatius" appears in the same fragment as the word "leucòcits".<br><br>Do not consider any "+" or "-" that is not directly related to leukocytes.<br><br>Text:<br>{t}<br>Respond only with a JSON using this exact structure:<br>{{<br>"leucocits": true/false,<br>"reftext_leucocits": "..."<br>}}<br><br>Rules:<br>- If "leucocits" is false, leave "reftext" completely empty ("").<br>- If "leucocits" is true, include only the exact fragment of the text where positive leukocytes appear.<br>- Do not invent or add words.<br>- Do not add any text outside the JSON.</td>
<td>Analitza el següent text de pacient i indica si els leucòcits són positius o negatius.<br>Només considera positius si apareix la paraula "leucòcits" (o "leucocits:" ) al mateix fragment que: "Traçes", "+", "++" o "+++".<br>Només considera negatius si apareix "Negatiu" o "Negatius" al mateix fragment que la paraula "leucòcits".<br><br>No consideris cap "+" o "-" que no estigui relacionat directament amb leucòcits.<br><br>Text:<br>{t}<br><br>Respon només amb un JSON amb aquesta estructura exacta:<br><br>{{<br>"leucocits": true/false,<br>"reftext_leucocits": "..."<br>}}<br><br>Regles:<br>- Si "leucocits" és false, deixa "reftext" completament buit ("").<br>- Si "leucocits" és true, posa només el fragment exacte del text on apareixen els leucòcits positius.<br>- No inventis ni afegeixis paraules.<br>- No afegeixis cap text fora del JSON.</td>
<td>leuc[oò]cits?\s*:?\s*tra[çc]es|leuc[oò]cits?\s*:?\s*\+{1,3}|leuc[oò]cits?\s*:?\s*negatiu|leuc[oò]cits?\s*:?\s*negatius|leucocitos?\s*:?\s*tra[z]as|leucocitos?\s*:?\s*\+{1,3}|leucocitos?\s*:?\s*negativo|leucocitos?\s*:?\s*negativos|leucos\s*:?\s*tra[çc]es|leucos\s*:?\s*\+{1,3}</td>
</tr>
</table>

<table>
<tr>
<td>Lumbar Pain</td>
<td>PPL +, PPL dret +, PPL esquerra +, PPL dubtosa, PPL+/-, Lumbalgia, Lumbago, dolor de ronyons o dolor dorsolumbar</td>
<td>Analyse the following patient text and indicate whether lumbar pain is present.<br>Consider lumbar pain present if the text contains: "PPL +", "PPL dret +", "PPL esquerra +", "PPL dubtosa", "PPL+/-", "Lumbalgia", "Lumbago", "dolor de ronyons", or "dolor dorsolumbar".<br><br>Text:<br>{t}<br>Respond only with a JSON using this exact structure:<br>{{<br>"dolor_lumbar": true/false,<br>"reftext_dolorlumbar": "..."<br>}}<br><br>Rules:<br>- If "dolor_lumbar" is false, leave "reftext" completely empty ("").<br>- If "dolor_lumbar" is true, include in "reftext" only the exact fragment of the text that indicates lumbar pain.<br>- Do not invent or add words.<br>- Do not add any text outside the JSON.</td>
<td>Analitza el següent text de pacient i indica si hi ha dolor lumbar.<br>Considera que hi ha dolor lumbar si el text conté: "PPL +", "PPL dret +", "PPL esquerra +", "PPL dubtosa", "PPL+/-", "Lumbalgia", "Lumbago", "dolor de ronyons" o "dolor dorsolumbar".<br><br>Text:<br>{t}<br><br>Respon només amb un JSON amb aquesta estructura exacta:<br>{{<br>"dolor_lumbar": true/false,<br>"reftext_dolorlumbar": "..."  # Inclou només el text exacte que indica dolor lumbar; deixa buit si dolor_lumbar=false.<br>}}<br><br>Regles:<br>- Si "dolor_lumbar" és false, deixa "reftext" completament buit ("").<br>- Si "dolor_lumbar" és true, posa a "reftext" només el fragment exacte del text que indica dolor lumbar.<br>- No inventis ni afegeixis paraules.<br>- No afegeixis cap text fora del JSON.</td>
<td>ppl\s*:?\s*\+|ppl dret\s*:?\s*\+|ppl esquerra\s*:?\s*\+|<br>ppl dubtosa|ppl\s*\+?/-<br>\s*|lumbalgia|lumbago|<br>dolor de ronyons|dolor dorsolumbar|<br>dolor lumbar|dolor en la zona lumbar|<br>lumbar pain|kidney pain|renal pain</td>
</tr>
</table>

| | | | | |
|---|---|---|---|---|
| **Abdominal Pain** | dolor en l'abdomen, dolor abdominal, dolor en hipogastri o mal de panxa | Analyse the following patient text and indicate whether abdominal pain is present. Consider abdominal pain present if the text contains: "dolor en l'abdomen", "dolor abdominal", "dolor en hipogastri", or "mal de panxa".<br><br>Text:<br>{t}<br>Respond only with a JSON using this exact structure:<br>{{<br>"dolor_abdominal": true/false,<br>"reftext_dolorabdominal": "..."<br>}}<br><br>Rules:<br>- If "dolor_abdominal" is false, leave "reftext" completely empty ("").<br>- If "dolor_abdominal" is true, include in "reftext" only the exact fragment of the text that indicates abdominal pain.<br>- Do not invent or add words.<br>- Do not add any text outside the JSON. | Analitza el següent text de pacient i indica si hi ha dolor abdominal.<br>Considera que hi ha dolor abdominal si el text conté: "dolor en l'abdomen", "dolor abdominal", "dolor en hipogastri" o "mal de panxa".<br><br>Text:<br>{t}<br><br>Respon només amb un JSON amb aquesta estructura exacta:<br><br>{{<br>"dolor_abdominal": true/false,<br>"reftext_dolorabdominal": "..." # Inclou només el text exacte que indica dolor abdominal; deixa buit si dolor_abdominal=false.<br>}}<br><br>Regles:<br>- Si "dolor_abdominal" és false, deixa "reftext" completament buit ("").<br>- Si "dolor_abdominal" és true, posa a "reftext" només el fragment exacte del text que indica dolor abdominal.<br>- No inventis ni afegeixis paraules.<br>- No afegeixis cap text fora del JSON. | dolor en l'abdomen\|dolor abdominal\|dolor en hipogastri\|mal de panxa\|<br>dolor de panxa\|dolor en el ventre\|dolor en la zona abdominal\|<br>abdominal pain\|stomach pain\|belly pain\|pain in the abdomen |

<table>
<tr><td>Haematuria</td><td>sang en orina, orina vermella, orina hemàtica, orina sanguinolenta o orina fosca</td><td>Analyse the following patient text and indicate whether haematuria is present.<br>Consider haematuria present if the text contains: "sang en orina", "orina vermella", "orina hemàtica", "orina sanguinolenta", or "orina fosca".<br><br>Text:<br>{t}<br>Respond only with a JSON using this exact structure:<br>{{<br>"hematuria": true/false,<br>"reftext_hematuries": "..."<br>}}<br><br>Rules:<br>- If "hematuria" is false, leave "reftext" completely empty ("").<br>- If "hematuria" is true, include in "reftext" only the exact fragment of the text that indicates haematuria.<br>- Do not invent or add words.<br>- Do not add any text outside the JSON.</td><td>Analitza el següent text de pacient i indica si hi ha hematúria.<br>Considera que hi ha hematúria si el text conté: "sang en orina", "orina vermella", "orina hemàtica", "orina sanguinolenta" o "orina fosca".<br><br>Text:<br>{t}<br><br>Respon només amb un JSON amb aquesta estructura exacta:<br><br>{{<br>"hematuria": true/false,<br>"reftext_hematuries": "..." # Inclou només el text exacte que indica hematúria; deixa buit si hematuria=false.<br>}}<br><br>Regles:<br>- Si "hematuria" és false, deixa "reftext" completament buit ("").<br>- Si "hematuria" és true, posa a "reftext" només el fragment exacte del text que indica hematúria.<br>- No inventis ni afegeixis paraules.<br>- No afegeixis cap text fora del JSON.</td><td>sang(re)?\\s*:?\\s*\\++|hematies\\s*:?\\s*\\++|hties\\s*:?\\s*\\++|htes\\s*:?\\s*\\++|<br><br>hemat\\s*:?\\s*\\++|hematuries\\s*:?\\s*\\++|hemat\\?ries\\s*:?\\s*\\++|orina vermella|<br>orina hem\\?tica|orina fosca|orina sanguinolenta|hemturia\\s*:?\\s*\\++|hematurias?\\s*:?\\s*\\++|<br>sang positiu|sang(re)? en orina|hemat\\?es\\s*:?\\s*\\++|orina hematurica|orina sang|sang(re)? en la orina|orinas oscuras</td></tr>
</table>

**Supplementary Table 2. Performance metrics from gold standard EHR MEAP narratives in cases and controls.**

| | TP | TN | FP | FN | Apparent Prevalence | True Prevalence | Sensitivity | Specificity | Accuracy | PPV | NPV |
|---|---|---|---|---|---|---|---|---|---|---|---|
| **CASES (N=30)** | 33 | 122 | 5 | 7 | 22.8 [ 16.6 , 29.9 ] | 24.0 [ 17.7 , 31.2 ] | 82.5 [ 67.2 , 92.7 ] | 96.1 [ 91.1 , 98.7 ] | 92.8 [ 87.8 , 96.2 ] | 86.8 [ 71.9 , 95.6 ] | 94.6 [ 89.1 , 97.8 ] |
| **Clinical feature, n (%)** | | | | | | | | | | | |
| **Fever** | 4 | 24 | 0 | 2 | 13.3 [ 3.8 , 30.7 ] | 20.0 [ 7.7 , 38.6 ] | 66.7 [ 22.3 , 95.7 ] | 100.0 [ 85.8 , 100.0 ] | 93.3 [ 77.9 , 99.2 ] | 100.0 [ 39.8 , 100.0 ] | 92.3 [ 74.9 , 99.1 ] |
| **Nitrites** | 2 | 23 | 0 | 0 | 8.0 [ 1.0 , 26.0 ] | 8.0 [ 1.0 , 26.0 ] | 100.0 [ 15.8 , 100.0 ] | 100.0 [ 85.2 , 100.0 ] | 100.0 [ 86.3 , 100.0 ] | 100.0 [ 15.8 , 100.0 ] | 100.0 [ 85.2 , 100.0 ] |
| **Leukocites** | 5 | 17 | 1 | 3 | 23.1 [ 9.0 , 43.6 ] | 30.8 [ 14.3 , 51.8 ] | 62.5 [ 24.5 , 91.5 ] | 94.4 [ 72.7 , 99.9 ] | 84.6 [ 65.1 , 95.6 ] | 83.3 [ 35.9 , 99.6 ] | 85.0 [ 62.1 , 96.8 ] |
| **Lumbar Pain** | 5 | 24 | 0 | 1 | 16.7 [ 5.6 , 34.7 ] | 20.0 [ 7.7 , 38.6 ] | 83.3 [ 35.9 , 99.6 ] | 100.0 [ 85.8 , 100.0 ] | 96.7 [ 82.8 , 99.9 ] | 100.0 [ 47.8 , 100.0 ] | 96.0 [ 79.6 , 99.9 ] |
| **Abdominal Pain** | 6 | 20 | 0 | 1 | 22.2 [ 8.6 , 42.3 ] | 25.9 [ 11.1 , 46.3 ] | 85.7 [ 42.1 , 99.6 ] | 100.0 [ 83.2 , 100.0 ] | 96.3 [ 81.0 , 99.9 ] | 100.0 [ 54.1 , 100.0 ] | 95.2 [ 76.2 , 99.9 ] |
| **Haematuria** | 11 | 14 | 4 | 0 | 51.7 [ 32.5 , 70.6 ] | 37.9 [ 20.7 , 57.7 ] | 100.0 [ 71.5 , 100.0 ] | 77.8 [ 52.4 , 93.6 ] | 86.2 [ 68.3 , 96.1 ] | 73.3 [ 44.9 , 92.2 ] | 100.0 [ 76.8 , 100.0 ] |
| **CONTROLS (N=30)** | 28 | 133 | 4 | 5 | 18.8 [ 13.2 , 25.5 ] | 19.4 [ 13.8 , 26.2 ] | 84.8 [ 68.1 , 94.9 ] | 97.1 [ 92.7 , 99.2 ] | 94.7 [ 90.2 , 97.6 ] | 87.5 [ 71.1 , 96.5 ] | 96.4 [ 91.7 , 98.8 ] |
| **Symptoms, n (%)** | | | | | | | | | | | |
| **Fever** | 1 | 27 | 0 | 2 | 3.3 [ 0.1 , 17.2 ] | 10.0 [ 2.1 , 26.5 ] | 33.3 [ 0.8 , 90.6 ] | 100.0 [ 87.2 , 100.0 ] | 93.3 [ 77.9 , 99.2 ] | 100.0 [ 2.5 , 100.0 ] | 93.1 [ 77.2 , 99.2 ] |
| **Nitrites** | 5 | 20 | 1 | 0 | 23.1 [ 9.0 , 43.6 ] | 19.2 [ 6.6 , 39.4 ] | 100.0 [ 47.8 , 100 .0] | 95.2 [ 76.2 , 99.9 ] | 96.2 [ 80.4 , 99.9 ] | 83.3 [ 35.9 , 99.6 ] | 100.0 [ 83.2 , 100.0 ] |
| **Leukocites** | 9 | 16 | 0 | 0 | 36.0 [ 18.0 , 57.5 ] | 36.0 [ 18.0 , 57.5 ] | 100.0 [ 66.4 , 100.0 ] | 100.0 [ 79.4 , 100.0 ] | 100.0 [ 86.3 , 100.0 ] | 100.0 [ 66.4 , 100.0 ] | 100.0 [ 79.4 , 100.0 ] |
| **Lumbar Pain** | 2 | 26 | 0 | 2 | 6.7 [ 0.8 , 22.1 ] | 13.3 [ 3.8 , 30.7 ] | 50.0 [ 6.8 , 93.2 ] | 100.0 [ 86.8 , 100.0 ] | 93.3 [ 77.9 , 99.2 ] | 100.0 [ 15.8 , 100.0 ] | 92.9 [ 76.5 , 99.1 ] |
| **Abdominal Pain** | 2 | 27 | 0 | 1 | 6.7 [ 0.8 , 22.1 ] | 10.0 [ 2.1 , 26.5 ] | 66.7 [ 9.4 , 99.2 ] | 100.0 [ 87.2 , 100.0 ] | 96.7 [ 82.8 , 99.9 ] | 100.0 [ 15.8 , 100.0 ] | 96.4 [ 81.7 , 99.9 ] |
| **Haematuria** | 9 | 17 | 3 | 0 | 41.4 [ 23.5 , 61.1 ] | 31.0 [ 15.3 , 50.8 ] | 100.0 [ 66.4 , 100.0 ] | 85.0 [ 62.1 , 96.8 ] | 89.7 [ 72.6 , 97.8 ] | 75.0 [ 42.8 , 94.5 ] | 100.0 [ 80.5 , 100.0 ] |

*TP: True Positive, TN: True Negative, FP: False Positive, FN: False Negative, PPV/NPV: Positive/Negative Predictive Value*

### Supplementary Table 3. Prompts to create synthetic clinical free-texts

| LLM | Prompt (English) |
|---|---|
| **Grok 4.1** | TASK: Create a synthetic table with free text for 60 patients.<br><br>CONTEXT:<br>I want you to create an Excel file with 60 cases/patients. In total, I want 30 positive (they have at least one of the symptoms) and 30 negative (they do not have the symptom) cases for each of the symptoms I will list below. I want you to invent an 'id' column with a patient identifier and a 'text' column where you write free text as if you were a doctor or nurse, containing the characteristics of the symptoms.<br><br>The symptoms are 6: fever, nitrites, leukocytes, lumbar pain, abdominal pain, and haematuria. Each case may have more than one symptom, but in total there must be 30 positive and 30 negative cases for each symptom.<br><br>Important details:<br>We consider fever: low-grade fever, temperature (Ta, Tª) above 37.5ºC, chills, dysthermic sensation, febrile episode, pyrexia, febrile syndrome, feeling feverish.<br><br>The symptom nitrites: can be Positive, Negative, +, -.<br>The symptom leukocytes: can be found as Traces, Negative, +, ++, +++.<br>The symptom lumbar pain: can be found as lumbar pain PPL +, PPL right/left +, PPL doubtful. PPL+/-.<br>The symptom abdominal pain: can be abdominal pain, pain in the hypogastrium, or stomach ache.<br><br>Finally haematuria: can appear as blood in urine, red urine, haematuric urine, sanguineous urine, or dark urine.<br><br>Objective: This table will be used to validate the phi4-mini model from the Ollama LLM. I am interested in having complex clinical profiles that could confuse the model.<br><br>Output: An Excel table with the fields id, text, and a logical column (TRUE/FALSE) for each symptom." |

**GPT 4.1**

I want you to create an Excel file with 60 cases/patients. In total, I want 30 positive and 30 negative cases for each of the symptoms I will list below. I want you to invent an 'id' column with a patient identifier and a 'text' column where you write a text as if you were a doctor, containing the characteristics of the symptoms.

The symptoms are: fever, nitrites, leukocytes, lumbar pain, abdominal pain, and haematuria.

We consider fever to also include: low-grade fever, temperature (Ta, Tª) above 37.5ºC, chills, dysthermic sensation, febrile episode, pyrexia, febrile syndrome, feeling feverish.

The symptom nitrites can be Positive, Negative, +, -.

The symptom leukocytes can also be found as Traces, Negative, +, ++, +++.

The symptom lumbar pain can also be found as lumbar pain PPL +, PPL right/left +, PPL doubtful. PPL+/-.

The symptom abdominal pain can also be abdominal pain, pain in the hypogastrium, or stomach ache.

And finally haematuria, which can also appear as blood in urine, red urine, haematuric urine, sanguineous urine, or dark urine.

Each case may have more than one symptom, but in total there must be 30 positive and 30 negative cases for each symptom. I want to use the text to validate the phi4-mini model from Ollama AI, so try to create complex texts that could confuse the model.

---

**Supplemental Table 4. Cohen's kappa of agreement between Phi4 and Gemma 3:4b**

| Gold standard MEAP narratives (N=60) | Cohen's kappa [95% CI] |
|---|---|
| **Has at least one of the features, n (%)** | 66.9 [47.4, 86.4] |
| **Clinical features, n (%)** | |
| **Fever** | 35 [11.2, 58.8] |
| **Nitrites** | 61.3 [37.7, 84.9] |
| **Leukocytes** | 79.3 [63.7, 94.8] |
| **Lumbar Pain** | 66.3 [45.5, 87.2] |
| **Abdominal Pain** | 52.4 [30.4, 74.3] |
| **Haematuria** | 53.9 [33.5, 74.4] |

**Supplemental Table 5. Performance metrics from gold standard EHR MEAP narratives using Gemma3:4b SML.**

| | TP | TN | FP | FN | Apparent Prev. | True Prev. | Sensitivity | Specificity | Accuracy | PPV | NPV |
|---|---|---|---|---|---|---|---|---|---|---|---|
| **Has at least one of the features, n (%)** | 71 | 215 | 49 | 2 | 35.6 [ 30.5 , 41.0 ] | 21.7 [ 17.4 , 26.4 ] | 97.3 [ 90.5 , 99.7 ] | 81.4 [ 76.2 , 85.9 ] | 84.9 [ 80.6 , 88.5 ] | 59.2 [ 49.8 , 68 ] | 99.1 [ 96.7 , 99.9 ] |
| **Clinical features, n (%)** | | | | | | | | | | | |
| **Fever** | 8 | 41 | 10 | 1 | 30.0 [ 18.8 , 43.2 ] | 15.0 [ 7.1 , 26.6 ] | 88.9 [ 51.8 , 99.7 ] | 80.4 [ 66.9 , 90.2 ] | 81.7 [ 69.6 , 90.5 ] | 44.4 [ 21.5 , 69.2 ] | 97.6 [ 87.4 , 99.9 ] |
| **Nitrites** | 6 | 38 | 6 | 1 | 23.5 [ 12.8 , 37.5 ] | 13.7 [ 5.7 , 26.3 ] | 85.7 [ 42.1 , 99.6 ] | 86.4 [ 72.6 , 94.8 ] | 86.3 [ 73.7 , 94.3 ] | 50.0 [ 21.1 , 78.9 ] | 97.4 [ 86.5 , 99.9 ] |
| **Leukocytes** | 17 | 33 | 1 | 0 | 35.3 [ 22.4 , 49.9 ] | 33.3 [ 20.8 , 47.9 ] | 100.0 [ 80.5 , 100.0 ] | 97.1 [ 84.7 , 99.9 ] | 98.0 [ 89.6 , 100.0 ] | 94.4 [ 72.7 , 99.9 ] | 100.0 [ 89.4 , 100.0 ] |
| **Lumbar Pain** | 10 | 41 | 9 | 0 | 31.7 [ 20.3 , 45.0 ] | 16.7 [ 8.3 , 28.5 ] | 100.0 [ 69.2 , 100.0 ] | 82.0 [ 68.6 , 91.4 ] | 85.0 [ 73.4 , 92.9 ] | 52.6 [ 28.9 , 75.6 ] | 100.0 [ 91.4 , 100.0 ] |
| **Abdominal Pain** | 10 | 38 | 9 | 0 | 33.3 [ 21.4 , 47.1 ] | 17.5 [ 8.7 , 29.9 ] | 100.0 [ 69.2 , 100.0 ] | 80.9 [ 66.7 , 90.9 ] | 84.2 [ 72.1 , 92.5 ] | 52.6 [ 28.9 , 75.6 ] | 100.0 [ 90.7 , 100.0 ] |
| **Haematuria** | 20 | 24 | 14 | 0 | 58.6 [ 44.9 , 71.4 ] | 34.5 [ 22.5 , 48.1 ] | 100.0 [ 83.2 , 100.0 ] | 63.2 [ 46.0 , 78.2 ] | 75.9 [ 62.8 , 86.1 ] | 58.8 [ 40.7 , 75.4 ] | 100.0 [ 85.8 , 100.0 ] |

*TP: True Positive, TN: True Negative, FP: False Positive, FN: False Negative, PPV/NPV: Positive/Negative Predictive Value*
